# Cost-effective and Flexible Preimplantation Genetic Testing (PGT) Using Adaptive Sampling-based Targeted Nanopore Sequencing (ASTN-seq)

**DOI:** 10.1101/2025.01.03.24319826

**Authors:** Zhiqiang Zhang, Shujing He, Taoli Ding, Xiaoyan Liang, Cong Fang, Haitao Zeng, Linan Xu, Xiaolan Li, Lei Jia, Shihui Zhang, Wenlong Su, Peng Sun, Ji Yang, Jun Ren, Sijia Lu, Zi Ren

**Affiliations:** Reproductive Medicine Center, The Sixth Affiliated Hospital, Sun Yat-sen University, Guangzhou 510655, China; Guangdong Engineering Technology Research Center of Fertility Preservation, Guangzhou 510655, China; Biomedical Innovation Center, The Sixth Affiliated Hospital, Sun Yat-sen University, Guangzhou 510655, China; Yikon Genomics Company, Ltd., Suzhou 215000, China

**Keywords:** Third-generation sequencing (TGS), Region of interest (ROI), Monogenic diseases, Chromosomal disorders, Haplotype linkage analysis

## Abstract

Genetic diseases encompass a spectrum of disorders resulting from DNA variations. Preimplantation genetic testing (PGT) is a critical strategy for preventing recurrent miscarriage, foetal malformations, and the birth of children affected by chromosomal abnormalities and monogenic disorders. Traditional PGT techniques necessitate comprehensive pedigree genetic data for haplotype linkage analysis, whereas PGT employing third-generation sequencing (TGS) has distinct advantages, particularly in cases of incomplete pedigree information, *de novo* mutations, and complex pathogenic variants. Nevertheless, the widespread application of TGS-based PGT in clinical practice encounters hurdles owing to its high costs. Targeted sequencing technologies present a promising solution by selectively enriching regions of interest while disregarding nontargeted areas, offering a more cost-effective and flexible alternative. In this study, we employed next-generation sequencing (NGS) and adaptive sampling-based targeted nanopore sequencing (ASTN-seq) to analyse samples from five couples who carried balanced translocations and *HBB* gene pathogenic mutations, as well as three additional couples with monogenic diseases caused by mutations in *PKD1*, *ASNS*, or *ALPL*. ASTN-seq successfully identified various mutations and facilitated haplotype linkage analysis, confirming its accuracy and reliability. Successful embryo transfer and subsequent prenatal diagnosis in certain families underscore the potential of ASTN-seq in assisted reproduction. Compared with traditional NGS-based PGT techniques, our work highlights that ASTN-seq is a promising tool for PGT, offering cost-effective solutions for PGT, especially for incomplete pedigrees and *de novo* mutations.

## Introduction

Genetic diseases mainly encompass disorders resulting from chromosomal abnormalities and monogenic variants [1]. Chromosomal disorders, such as reciprocal translocations, are significant causes of recurrent miscarriage and foetal malformations [2]. Couples with balanced reciprocal translocations have a 50% chance of having recurrent pregnancy loss and a 20% risk of having children with unbalanced chromosomal rearrangements [3]. Monogenic diseases are hereditary disorders stemming from mutations in specific genes. Thalassemia, including α- and β-thalassemia, is one of the five major monogenic diseases of humans worldwide [4]. In mainland China, the overall prevalence of α-thalassemia, β-thalassemia, and α+β-thalassemia were reported to be 7.88%, 2.21%, and 0.48% respectively [5]. Given the limited treatment options for most genetic diseases, preventive measures such as preimplantation genetic testing for chromosomal structural rearrangements (PGT-SR) and preimplantation genetic testing for monogenic/single-gene diseases (PGT-M) are critically important [6].

PGT-SR and PGT-M are usually based on whole genome amplification (WGA) and next-generation sequencing (NGS) to construct haplotypes for distinguishing whether the embryo carries chromosomal structural rearrangements or pathogenic mutations [6]. These conventional approaches require comprehensive genetic information on pedigrees, which limits their applicability in instances of incomplete pedigrees or *de novo* mutations. Currently, conventional clinical approaches addressing these challenges involve embryo extrapolation and the single-sperm methodology. Nonetheless, embryo extrapolation hinges on the availability of a reference embryo, whereas the single-sperm technique is intricate, expensive, and exclusive to male patients. When confronted with intricate pathogenic mutations such as large-fragment structural variants (SVs), pseudogene interference, and dynamic mutations, conventional NGS-based methods exhibit limitations in embryo extrapolation and application in the single-sperm approach. In contrast, third-generation sequencing (TGS) has significant advantages over NGS [7–15] and has emerged as a superior alternative. The read length of TGS is long enough to include pathogenic variants and single nucleotide polymorphisms (SNPs) in the same sequencing reads, generating natural haplotypes. TGS-based preimplantation genetic testing (PGT) has been utilized to distinguish carriers from noncarrier embryos directed against balanced reciprocal translocations [7, 11–14], inversions [13], or various single-gene mutations [8, 11, 13, 15]. The breakpoints of balanced reciprocal translocation, inversion, and deletion can be precisely identified at single-base resolution [7–9, 12]. TGS offers new solutions for PGT, especially in cases of incomplete pedigrees and *de novo* mutations.

Despite the substantial potential of TGS, its application in the clinic faces economic challenges. TGS methods necessitate expensive flow cells, reagents, computing resources, and so on. The high costs pose challenges for many laboratories, medical institutions, and patients seeking to adopt them. Adaptive sampling-based targeted nanopore sequencing (ASTN-seq) technology optimizes the sequencing process by focusing on specific regions of interest (ROIs) in the genome, thereby saving sequencing time and lowering the cost of reagents, flow cells, and computing resources. Additionally, ASTN-seq ensures a high coverage depth of the ROI [16], effectively reducing unnecessary sequencing. Its efficient target sequence enrichment strategy delivers more accurate results, reducing the complexity of data analysis and further diminishing overall operating costs. These advantages maximize the potential of the nanopore sequencer, easing the burden of initial investment and enabling more laboratories and medical institutions to adopt the technology. For patients, ASTN-seq offers a significant cost advantage over TGS-based whole-genome sequencing (WGS), potentially reducing costs by 50% or more, thereby facilitating clinical adoption and benefiting patients. In the future, the cost of ASTN-seq-based PGT could equal that of NGS. Additionally, the ASTN-seq method is versatile, allowing targeted sequencing of various genomic regions and detection of multiple mutation types. This means that ASTN-seq is applicable for both PGT-M and PGT-SR or even both simultaneously. For patients, especially those who carry translocations and single-gene mutations at the same time, PGT is less expensive and simpler. Medical institutions can also streamline experimental procedures, reducing operational and management costs. The introduction of ASTN-seq represents an innovative solution to overcome the economic barriers associated with TGS implementation, making it a practical and economically viable option for PGT.

To establish the feasibility and effectiveness of ASTN-seq in assisted reproductive technology (ART), we applied this method to concurrently perform PGT-SR and PGT-M on five families affected by chromosomal rearrangement diseases and monogenic diseases. Additionally, PGT-M was conducted for three couples affected solely by monogenic diseases. The accuracy of ASTN-seq was further validated through prenatal diagnosis, to provide a valuable reference for the clinical implementation of this technology. Our work demonstrates that ASTN-seq is reliable and versatile, with potential applications in both PGT-M and PGT-SR. ASTN-seq effectively reduces the cost of TGS-based PGT, thereby benefiting clinical institutions and patients.

## Materials and methods

### Patients

Between February 2021 and October 2024, eight couples who underwent PGT-M and/or PGT-SR cycles at our reproductive medicine centre were enrolled in this study. The inclusion criteria for these couples were as follows: one partner carrying a balanced translocation, with both partners carrying *HBB* gene pathogenic mutations, or one or both partners carrying pathogenic mutations in genes other than the *HBB* gene. Detailed information about the eight families is provided in **Table 1**. The entire process is outlined in detail in Figure S1. Complete informed consent was obtained from all of the participating couples.

**Table 1.**
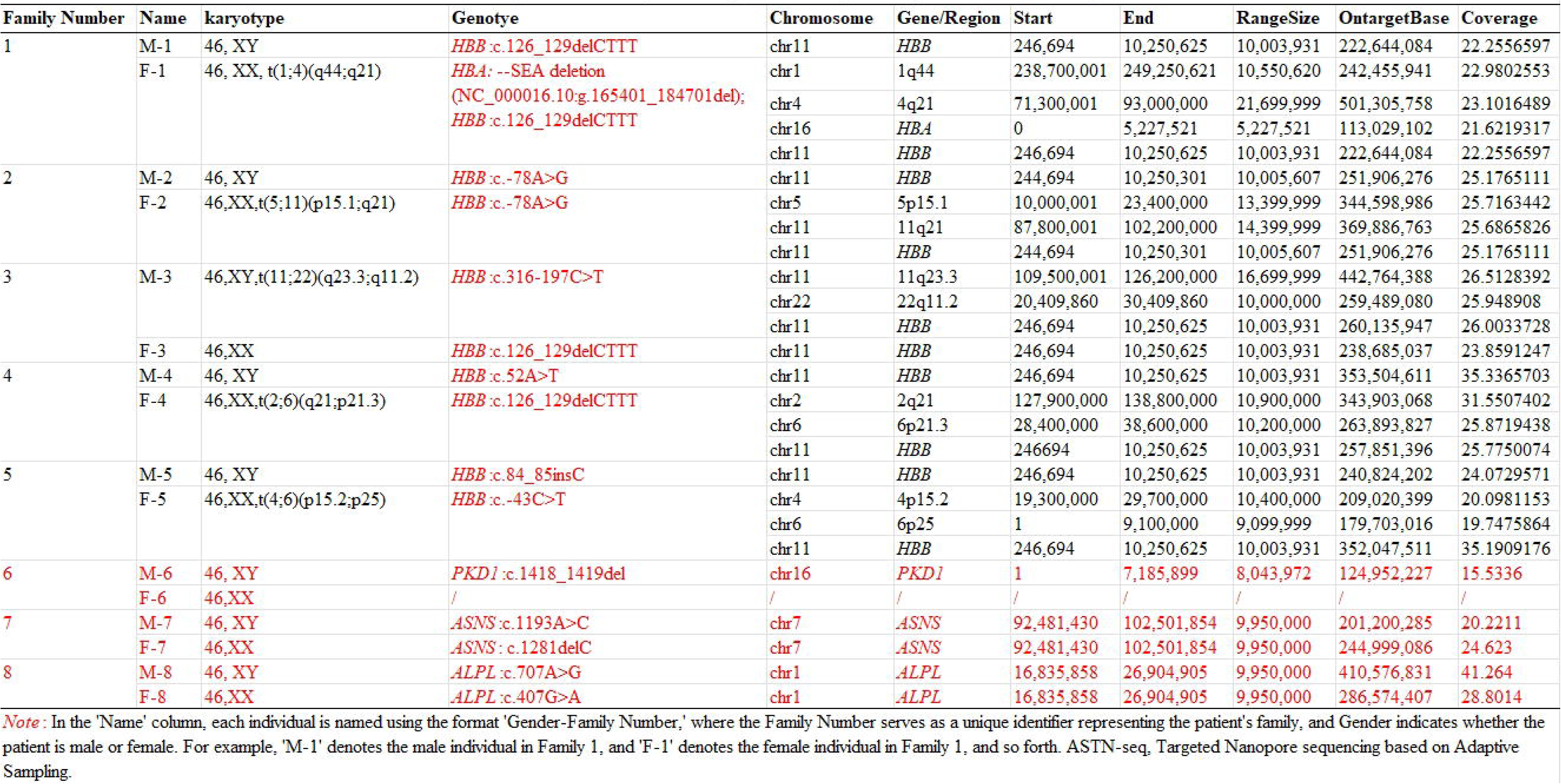
Genetic Profile of Families 1 to 8 and Comprehensive Statistics on ASTN-seq Sequencing Data for Individual Samples, Encompassing Target Region Commencement and Termination Points, Target Region Dimensions, ASTN-seq Data Volume, and Depth of the Targeted Region

### DNA extraction and library preparation

Peripheral blood was collected from the participating couples. High-molecular-weight genomic DNA was prepared using the SDS method and purified by using a QIAGEN® Genomic Kit (Cat# 13343, QIAGEN, Hilden, Germany) according to the instructions provided by the manufacturer. Agarose gels (1%) were used to assess the degradation and contamination of the extracted DNA. In addition, DNA purity was measured with a NanoDrop^TM^ One UV[Vis spectrophotometer (Thermo Fisher Scientific, Waltham, MA). For pure DNA, the OD 260/280 should range from 1.8 to 2.0, and the OD 260/230 should range from 2.0 to 2.2. The DNA concentration was measured with a Qubit® 3.0 fluorometer (Invitrogen, Carlsbad, CA).

For the preparation of the ONT libraries, 2 µg of DNA was used per sample. The size of long DNA fragments was selected via the BluePippin system (Sage Science, Beverly, MA). With the Ultra II End Repair/dA-tailing Kit (Cat# E7546, New England Biolabs, Ipswich, MA), the DNA fragment ends were repaired, and ligation reactions were performed. The adapter in the LSK109 kit (Oxford Nanopore Technologies, Oxford, UK) was used for further ligation, and a Qubit® 3.0 fluorometer was used to quantify the size of the library fragments.

### ASTN-seq

ASTN-seq is a targeted sequencing technology that leverages the distinctive asynchronous mode of nanopore sequencing. The methodology involves comparing the real-time tunneling current signals of DNA molecules traversing nanopores with the current signal pattern of the target sequence. If the signal comparison yields consistency, sequencing progresses. In cases of inconsistency, the nanopore sequencer applies a reverse voltage to expel the DNA molecule from the nanopore, allowing the continuation of sequencing other DNA molecules. A schematic diagram of ASTN-seq is shown in **Figure 1**.

**Figure 1.**
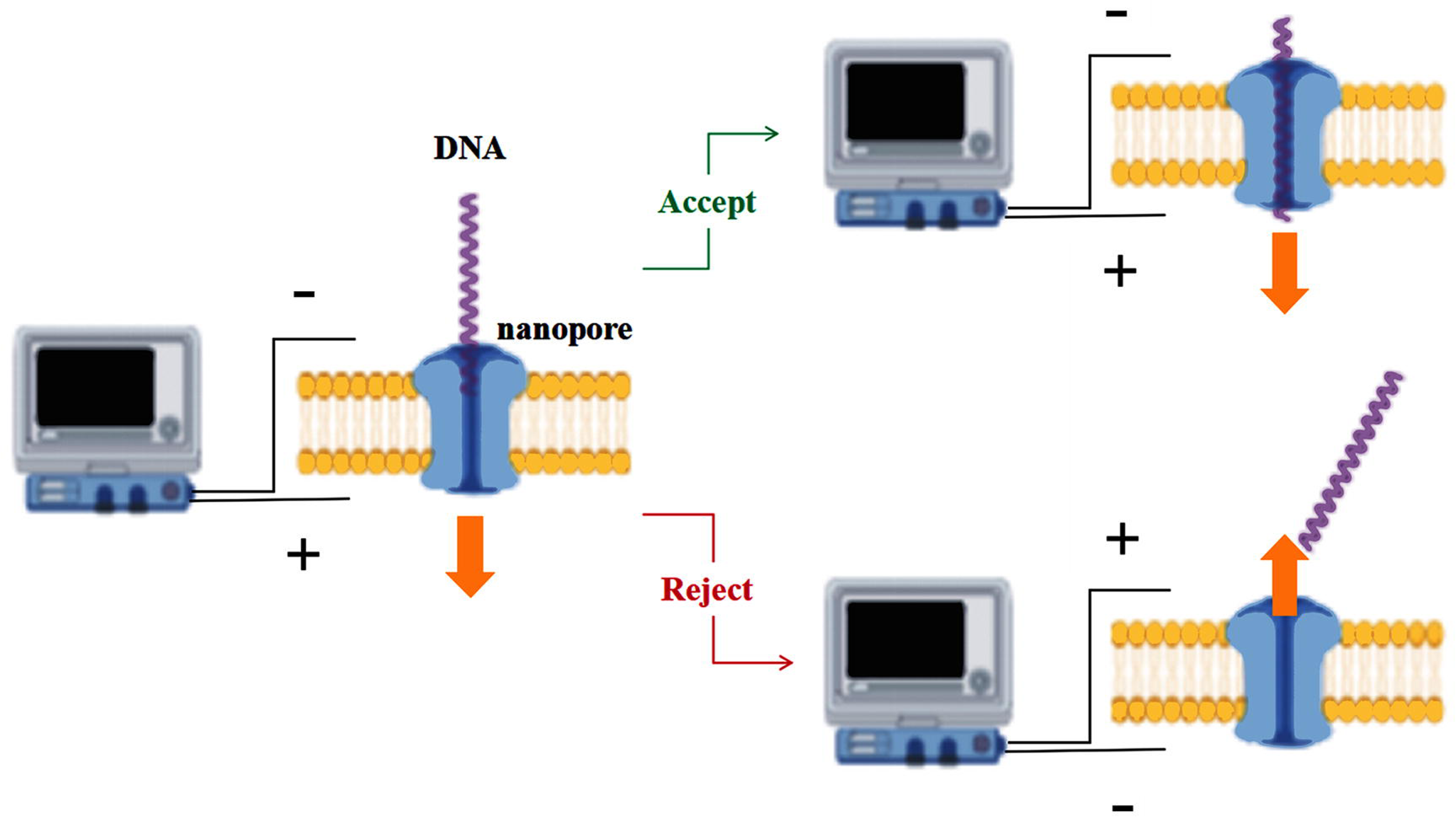
Schematic diagram of Nanopore-based Adaptive Sampling Targeted Nanopore Sequencing (ASTN-seq) ASTN-seq is a method of software-controlled enrichment unique to nanopore sequencing platforms. The yellow parts represent the lipid membrane, the blue parts represent the nanopore, and the purple curves represent the DNA molecules. When a DNA molecule passes through a nanopore, it generates real-time sequencing electrical signals. The sequencing software aligns these signals against a target sequence. If the alignment is consistent (accept), the DNA molecule being sequenced is our target sequence, and the sequencing process continues. If the alignment is inconsistent (reject), the sequencer applies a reverse voltage to expel the negatively charged DNA molecule from the nanopore.

The sequence was executed on a MinION sequencer (Oxford Nanopore Technologies, Oxford, UK). The sequencing instrument was linked to a server running Ubuntu 20.04, equipped with a GTX 3090 GPU. The third-party software readfish (https://github.com/LooseLab/readfish) was installed and utilized. It connects to the MinKNOW server in real time through the official Read Until API from Oxford Nanopore Technologies to perform targeted sequencing. Before initiating targeted sequencing, preparations should include reference sequences for alignment, readfish configuration files, and bed files containing target regions. Additionally, the parameter “break_reads_after_seconds=1.0” in the corresponding TOML configuration file located at /opt/ont/minknow/conf/package/sequencing should be modified to “break_reads_after_seconds=0.4”. After sequencing starts through the MinKNOW UI, readfish can be executed. Please note that readfish currently supports only Ubuntu systems with GPUs of sufficient performance. The installation of readfish depends on the MinKNOW and Guppy versions installed on the server and can be found at https://github.com/LooseLab/readfish.

In this study, the indicators used to evaluate the effectiveness of targeted sequencing included total data output, sequencing depth of the targeted region, enrichment efficiency, and enrichment factor. The total data output refers to the amount of data produced by a single MinION sequencing flow cell under the ASTN-seq operation. The enrichment efficiency is defined as the ratio of the data output from the targeted region to the total data output. The enrichment factor refers to the ratio of the sequencing depth of the targeted region to that of the nontargeted region.

### Embryo culture, blastocyst biopsy and WGA

All the collected oocytes were fertilized by intracytoplasmic sperm injection. The embryos were cultured individually in Vitrolife G-Series culture media (Vitrolife, Gothenburg, Sweden) in a 37 °C humidified atmosphere of 6% CO_2_ and 5% O_2_ balanced with N_2_. On day 5 or day 6, blastocysts were scored on the basis of the Gardner and Schoolcraft system [17]. Biopsy of blastocyst trophectoderm cells was performed as a routine procedure. Five to ten blastocyst trophoblast cells were removed and then transferred into RNase-DNase-free PCR tubes containing 5 mL of cell lysis buffer (Cat#XK043, Yikon Genomics, Suzhou, China). Biopsied cells underwent WGA by multiple annealing and looping-based amplification cycles for the detection of **copy number variants (**CNVs) and SNPs. After biopsy, the blastocysts were frozen by vitrification according to the manufacturer’s protocol (VT101, Kitazato, Fuji, Japan). In total, 45 embryos from eight families were obtained and detected.

### CNV detection and SNP genotyping

Following previously published procedures, genome-wide quantification of copy number was conducted based on the number of sequencing reads obtained through NGS. Furthermore, a high-throughput genotyping array, specifically the Infinium Asian Screening Array (Cat# 20016317, Illumina, San Diego, CA), was utilized to identify SNPs for each sample. This included genomic DNA extracted from peripheral blood and amplification products from embryonic biopsy cells generated through multiple annealing and looping-based amplification cycles, all of which were conducted following the manufacturer’s instructions. Only SNPs that were consistent between the chip and TGS were used for subsequent haplotype construction and linkage analysis.

### Mapping, detection of structural variants, phasing and haplotype linkage analysis

Raw data (raw reads) in fastq format were obtained from the electrical signal produced by MinION. In this process, Guppy basecalling software (v5.0.16) was used. To ensure the quality and reliability of the analysis, NanoFilt (v2.8.0, https://github.com/wdecoster/nanofilt) was used to remove low quality (Qphred<=7) and short reads (<1000 bp) from the raw data. The 50 bp bases in the head/tail were cut. We employed Minimap2(v2.26, https://github.com/lh3/minimap2) to align reads to reference genomes, including GRC37, GRCh38, and T2T-CHM13. The Minimap2 parameters used were -ax map-ont -L --MD -Y -t 20. SAMtools (v1.2, https://github.com/samtools/samtools) was subsequently utilized to convert the SAM-formatted files to BAM files, ensuring compatibility and facilitating downstream analyses. We analysed the BAM files using Sniffles (https://github.com/fritzsedlazeck/Sniffles), employing the parameters -t 12 --min_support 1 --num_reads_report -1. The initial results of structural variants were screened on the basis of high-quality variant reads and a karyotype diagnosis report. PEPPER-WhatsHap-DeepVariant (r0.7-gpu, https://github.com/kishwarshafin/pepper) was employed to report single-base mutations and InDels within samples, with simultaneous generation of haplotype results. By utilizing the BAM file as input and applying the specified parameters --ont_r9_guppy5_sup -g --phased_Output -t 12, a VCF file containing phasing information was generated. Python (v3.10.1) was then used to obtain the classification results of the target region. We applied a likelihood-based haplotyping approach, employing the hidden Markov model strategy, to discern the most likely haplotype configuration of embryos and other noprobands.

### Mutation validation and prenatal diagnosis

To validate the target mutations, PCR primers were designed using Primer3-Plus (http://primer3plus.com/). The designed primers were subsequently synthesized and utilized to amplify DNA extracted from embryonic biopsy cells via the conventional method. The expected amplicons were visualized on 2% ethidium bromide agarose gels, and the presence of the target regions was further confirmed by Sanger sequencing to detect the pathogenic mutation.

To evaluate foetal outcomes, we collected amniotic fluid samples from pregnant women. In the event of a miscarriage, we procure chorionic villus tissue postmiscarriage. The techniques for amniocentesis and karyotyping of amniotic fluid cells were conducted following established literature protocols [18]. For chorionic villus sampling, a sterile method was employed, and any attached blood was cleansed away with physiological saline. The villi were delicately isolated under an anatomical microscope, and 10 to 15 mg of this tissue was collected and finely chopped into a paste using ophthalmic scissors. This paste was then deposited into a sterile 15 mL centrifuge tube. To this tube, we added 2 mL of 0.25% trypsin-EDTA (GIBCO, Carlsbad, CA) and incubated the mixture at 37 °C with shaking for 30 minutes. The digestion process was terminated by adding 1 mL of calf serum (GIBCO, Carlsbad, CA), followed by centrifugation at 2000 rpm for 8 minutes. After the supernatant was discarded, approximately 1 mL of the pellet was retained, transferred to a 25-square centimetre cell culture flask (Thermo Scientific Nunc, Waltham, MA), and preloaded with 5 mL of AmnioMAX-II culture medium (GIBCO, Carlsbad, CA). This culture was maintained at 37 °C in an atmosphere containing 5% CO□ for three days. As soon as the colonies emerged, the medium was replenished, and the culture was continued for an additional 3 to 5 days. Three drops of colchicine were subsequently added to reach a final concentration of 20 μg/mL, and the mixture was incubated for another 2 to 3 hours before harvesting. The collection, processing, and G-banding analysis of the chorionic villus cells followed the same protocols as those utilized for amniotic fluid cells.

Gap-polymerase chain reaction and flow-through hybridization technology (Hybribio Limited, Hong Kong, China) were employed for the detection of foetal α-thalassemia mutations [19]. The carrier status of the foetal β-thalassemia gene was determined using the PCR reverse dot blot method [20]. Sanger sequencing was employed to verify mutations in the *PKD1*, *ASNS*, and *ALPL* genes. To determine the potential risk of maternal cell contamination in foetal tissue samples, such as chorionic villi or amniotic fluid, we employed quantitative fluorescence PCR to evaluate the presence of maternal cells within the samples. The application of this detection method strictly adhered to the detailed procedures outlined in the literature [21].

## Results

### Research and development of ASTN-seq

In this study, we investigated eight couples, and their detailed information is presented in Table 1. By utilizing karyotype results and genetic diagnosis reports, we identified target regions containing translocation breakpoints and single-gene pathogenic mutations. Taking F-1 from Family 1 as an example, her karyotype was 46, XX, t (1; 4) (q44; q21). Additionally, she was heterozygous for the --^SEA^ deletion (NC_000016.10: g.165401_184701del) in the *HBA1/2* gene and the *HBB*:c.126_129delCTTT mutation. Using the GRCh37 reference genome, we confirmed the targeted genomic regions as 1q44 (chr1: 238,700,001-249,250,621), 4q21 (chr4: 71,300,001-93,000,000), the *HBA* gene (chr16: 0-5,227,521), and the *HBB* gene (chr11: 246,694-10,250,625). The start and end positions, as well as the sizes of the target regions for all of the samples, are listed in Table 1.

ASTN-seq was performed on the DNA extracted from peripheral blood according to the target regions listed in Table 1. We subsequently evaluated the quality of the ASTN-seq data focusing on four aspects: enrichment efficiency, sequencing depth, fold enrichment, and read length distribution. In **Figure 2A**, the bar chart depicts the number of bases sequenced in the ROI and the total number of bases in each sample, whereas the line represents the enrichment efficiency of each sample. The mean and median values of enrichment efficiency were 6.96% and 6.69%, respectively. We also calculated the sequencing depth of the ROI and compared it with the depth of WGS under the same data output conditions, as shown in the box plot in **Figure 2B**. Under ASTN-seq, the mean and median sequencing depths of the target regions were 26.34× and 25.33×, respectively. Under WGS, the mean and median sequencing depths of the ROI were 3.26 and 2.99, respectively. A hypothesis test revealed a significant difference in sequencing depth between ASTN-seq and WGS (*P* = 3.86*10^-10^). The box plot in **Figure 2C** presents the summary statistics of the read length N50 of ASTN-seq in the ROI for each sample. Moreover, we defined fold enrichment as the ratio of sequencing depth in the ROI to that in the nontarget region. As shown in **Figure 2D**, the fold enrichment values of each sample obtained by ASTN-seq had mean and median values of 10.28 and 8.90, respectively, which were much higher than those obtained by WGS, with a theoretical value of 1.

**Figure 2.**
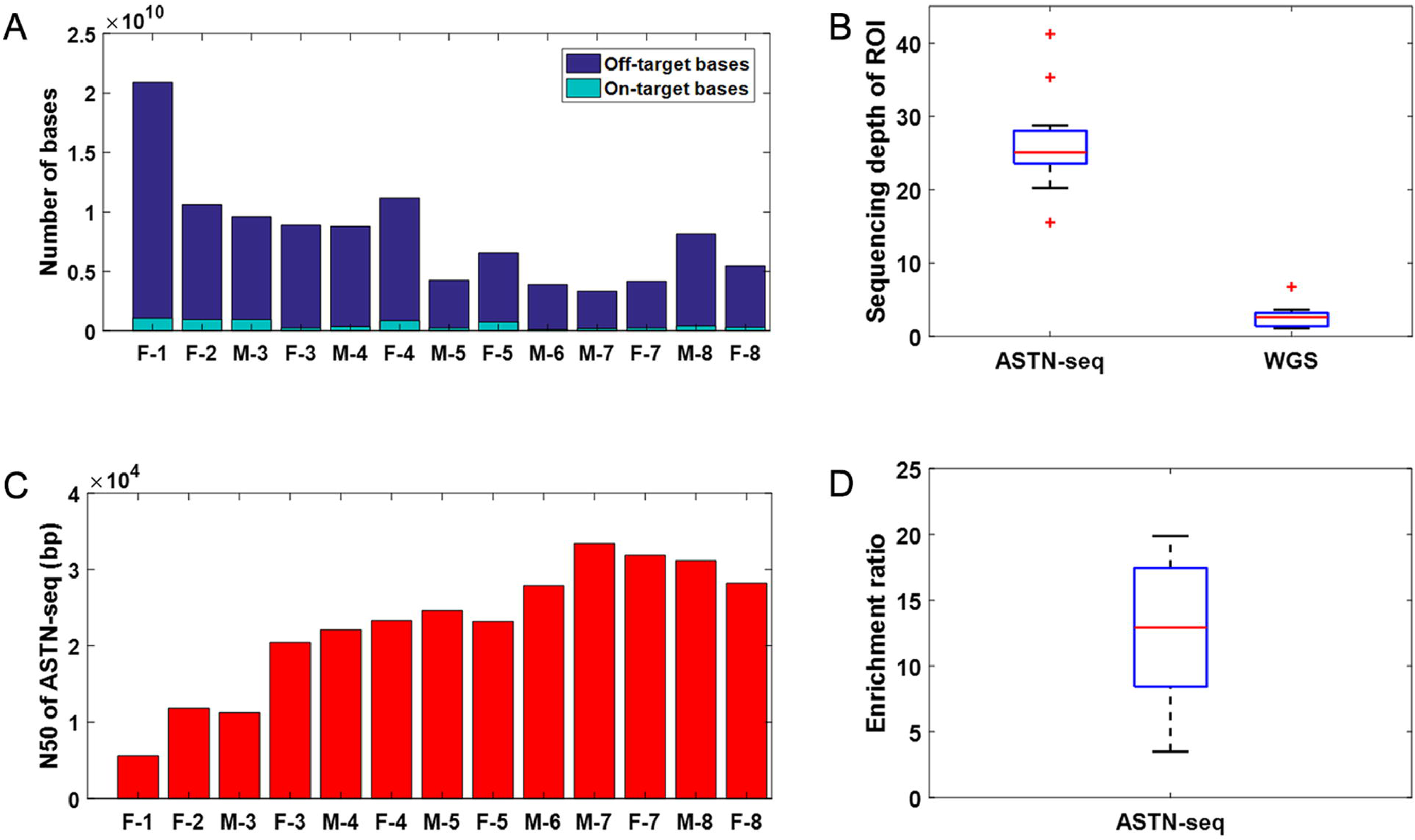
Statistical results of the sequencing data obtained through ASTN-seq. Panel (**A**) displays the counts of target and off-target bases. Panel (**B**) presents a boxplot demonstrating the depth of the target region for ASTN-seq compared with that for WGS with equivalent data volumes. Panel (**C**) depicts the read length N50 for all samples. Panel (**D**) shows the boxplot of enrichment ratios for ASTN-seq, with the enrichment ratio of WGS standardized to 1. ASTN-seq: nanopore-based adaptive sampling targeted nanopore sequencing; WGS: whole-genome sequencing; N50: 50% of the covered bases are found within contigs longer than this number. Each individual is labeled as ‘Gender-Family Number,’ e.g., ‘M-1’ for the male and ‘F-1’ for the female in Family 1, with the same format for others.

### Detection of various mutations and haplotype construction based on ASTN-seq

ASTN-seq can detect various types of mutations, including point mutations, large-fragment deletions, and translocations, with single-base resolution.

For F-1 in Family 1, the diagnostic report indicated the presence of a balanced translocation, a heterozygous --^SEA^ deletion (NC_000016.10: g.165401_184701del) in the *HBA1/2* gene, and a heterozygous *HBB*:c.126_129delCTTT mutation. M-1 only carries a heterozygous *HBB*:c.126_129delCTTT mutation. We performed ASTN-seq and bioinformatics analysis on this couple, successfully identifying the breakpoints associated with the balanced translocations as well as those linked to the thalassemia-associated mutations. Specifically, one breakpoint of the balanced translocation (1q44) is located at chr1:235414823, whereas the other (4q21) is located at chr4:66587123. ASTN-seq also successfully identified the heterozygous --^SEA^ deletion (NC_000016.10: g.165401_184701del) in the *HBA1/2* gene carried by F-1 and the heterozygous *HBB*:c.126_129delCTTT mutation carried by both individuals. The analysis results for this family are visually presented in an Integrative Genomics Viewer (IGV) plot in **Figure 3A**.

**Figure 3.**
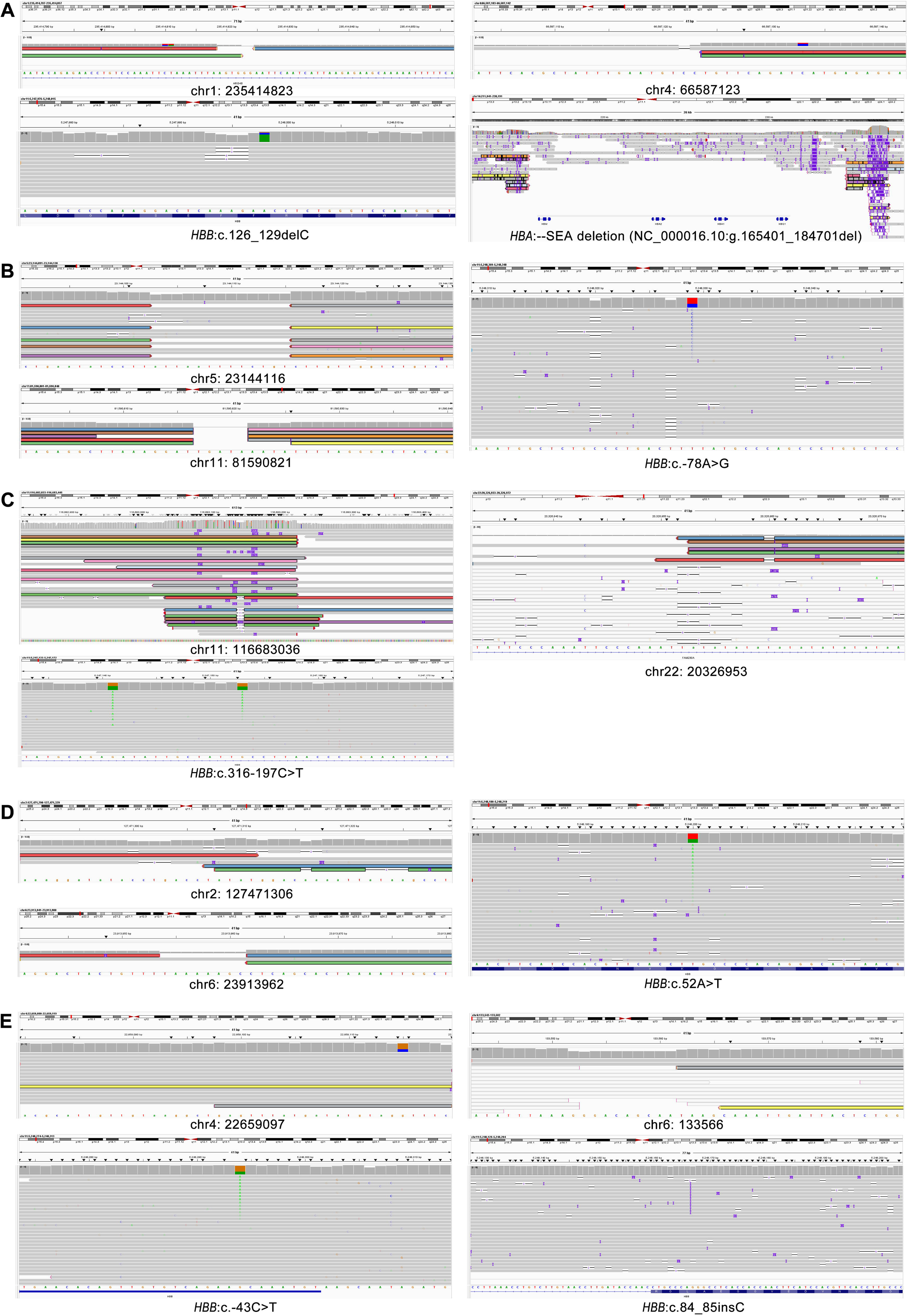
The IGVs of the variants. The horizontal bands represent sequencing reads. The coloured bands that are highlighted represent reads carrying pathogenic mutations. The grey bands represent those that do not carry translocation breakpoints or pathogenic variants. Panel (**A**) presents the IGV graphs of Family 1, which include the breakpoints of translocation, variants in the *HBB* gene, and the SEA deletion in the *HBA* gene. Panel (**B**) displays the IGV graphs of Family 2, featuring the breakpoints of translocation and a variant in the *HBB* gene. Panels (**C**), (**D**), and (**E**) show the IGV graphs of Family 3, Family 4, and Family 5 respectively. IGV: Integrative Genomics Viewer.

In Family 2, F-2 carries a balanced translocation with a karyotype of 46,XX,t(5;11)(p15.1;q21). Additionally, she harbour a heterozygous *HBB*:c.-78A>G mutation, whereas M-2 only possesses the heterozygous *HBB*:c.-78A>G mutation. The breakpoints for balanced translocation were validated at chr5:23144116 and chr11:81590821 through ASTN-seq. Moreover, ASTN-seq effectively identified the heterozygous *HBB*:c.-78A>G mutation carried by both individuals. The results for this family are visually depicted in the IGV plot shown in **Figure 3B**.

In Family 3, M-3 carries a balanced translocation with a karyotype of 46,XY,t(11;22)(q23.3;q11.2) and a heterozygous *HBB*:c.316-197C>T mutation. F-3 carries only the heterozygous *HBB*:c.126_129delCTTT mutation. Through ASTN-seq, we successfully identified the breakpoints of the balanced translocations and thalassemia-related mutations, as depicted in the IGV plot in **Figure 3C**. The translocation breakpoints are located at chr11:116683036 and chr22:20326953. The mutation sites in the *HBB* gene include the heterozygous *HBB*:c.316-197C>T mutation and the heterozygous *HBB*:c.126_129delCTTT mutation.

In Family 4, F-4 carries both a balanced translocation 46,XX,t(2;6)(q21;p21.3) and the heterozygous *HBB*:c.126_129delCTTT mutation, whereas M-4 only carries the heterozygous *HBB*:c.52A>T mutation. We conducted ASTN-seq for F-4 and M-4 to identify breakpoints of the balanced translocations and mutations associated with single-gene disorders. **Figure 3D** shows the translocation breakpoints on chromosomes 2q21 and 6p21.3 in F-4, which are located at chr2:127471306 and chr6:23913962, respectively. The *HBB*:c.52A>T mutation of M-4 is also depicted in Figure 3D. However, the *HBB* mutation *HBB*:c.126_129delCTTT of F-4 is not depicted again.

According to the diagnostic report, F-5 in Family 5 carries a balanced translocation 46,XX,t(4;6)(p15.2;p25) and a heterozygous *HBB*:c.-43C>T mutation, whereas M-5 carries only the heterozygous *HBB*:c.84_85insC mutation. **Figure 3E** illustrates the mutations detected by ASTN-seq in M-5 and F-5. On the basis of the analysis results, the specific location of the translocation breakpoint at 4p15.2 for F-5 was chr4:22659097, and the breakpoint at 6p25 was chr6:133566. The heterozygous *HBB*:c.-43C>T mutation and heterozygous *HBB*:c.84_85insC mutation are also shown in Figure 3E. Additionally, all translocation breakpoints identified by ASTN-seq were validated using PCR. With the exception of Family 3, the accuracy of the translocation breakpoints detected by ASTN-seq was confirmed through PCR verification in all other pedigrees (Figure S2). The failure of PCR validation in Family 3 is due to the high homology of the downstream region of the translocation breakpoint, which contains only three divergent sites, making primer design particularly challenging.

In addition to thalassemia, we also attempted PGT-M for other genetic diseases using ASTN-seq. In Family 6, M-6 carries a pathogenic mutation, *PKD1*:c.1418_1419del (p.Val473Alafs*45) for polycystic kidney disease, whereas F-6 is unaffected. In Family 7, both members of the couple carry gene mutations associated with an autosomal recessive (AR) genetic disorder. M-7 carries the mutation *ASNS*:c.1193A>C (p.Tyr398Ser), and F-7 carries the mutation *ASNS*:c.1281delC (p.Tyr428Ilefs*9). In Family 8, both members of the couple also carry mutations associated with an AR genetic disorder. M-8 carries the mutation *ALPL*:c.707A>G (p.Tyr236Cys), and F-8 carries the mutation *ALPL*:c.407G>A (p.Arg136His). All of the mutations mentioned in Families 6, 7, and 8 were successfully detected by ASTN-seq and subsequently verified by Sanger sequencing, as shown in Figure S3.

Furthermore, we conducted genotyping using SNPs from chips, WGS data from TGS, and ASTN-seq. The genotypes based on ASTN-seq, chips, and WGS data from TGS are consistent, demonstrating the accuracy and feasibility of the ASTN-seq-based haplotyping method.

### Detection of embryonic CNVs and haplotype linkage analysis

We retrieved multiple blastocysts from either day 5 or day 6 stages and isolated a limited number of trophectoderm cells. Following this, the biopsy cells underwent WGA, succeeded by the detection of CNVs and SNPs and the validation of mutations through Sanger sequencing. These comprehensive analyses enabled us to ascertain the euploidy status of the embryos and determine the presence of balanced translocations or pathogenic mutations. For the embryos from the eight pedigrees included in this study, we detected CNVs and performed haplotype linkage analysis using NGS as the standard. We subsequently performed haplotype linkage analysis on embryos via ASTN-seq to validate the feasibility and accuracy of ASTN-seq-based PGT.

The three embryos of Family 1 were designated E1-1-1, E1-1-2, and E1-1-3, among which E1-1-1 and E1-1-2 were euploid. The CNV results for these embryos are summarized in **Table 2**. Haplotype linkage analysis based on ASTN-seq was further conducted on E1-1-1 and E1-1-2. E1-1-1 only carried the maternal translocations and did not exhibit *HBA1/2* or *HBB* gene mutations inherited from the parents. E1-1-2, on the other hand, lacked the maternal translocation but harboured the paternal *HBB* gene mutation. The results of the haplotype linkage analysis are presented in **Figure 4**. The genotyping results were consistent with the results of NGS, confirming the accuracy and reliability of the ASTN-seq results. Sanger sequencing was also conducted to verify the embryos, and the results are presented in Figure S4A, further confirming the accuracy of the haplotype linkage analysis based on ASTN-seq.

**Table 2.**
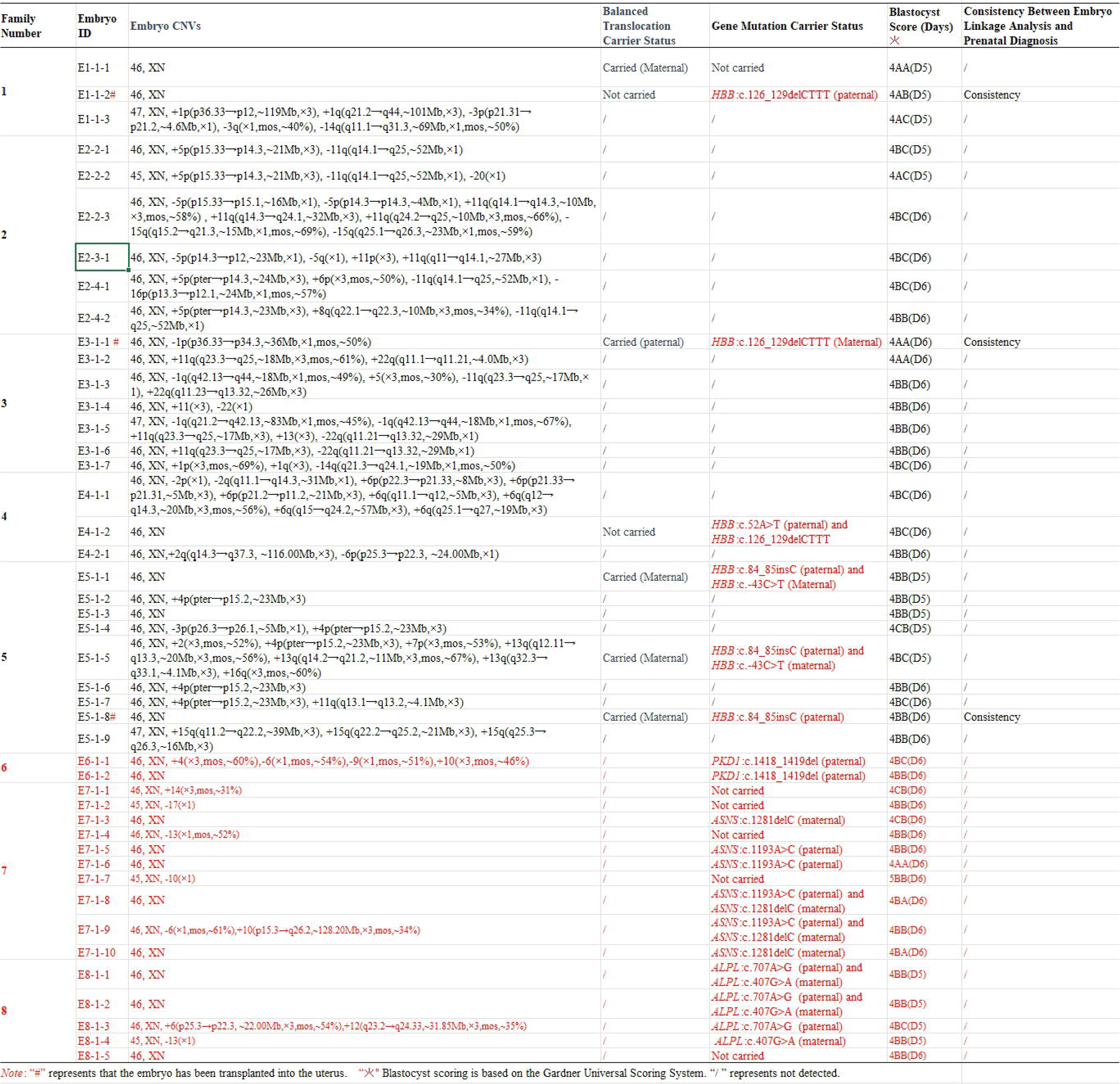
Embryo information table.

**Figure 4.**
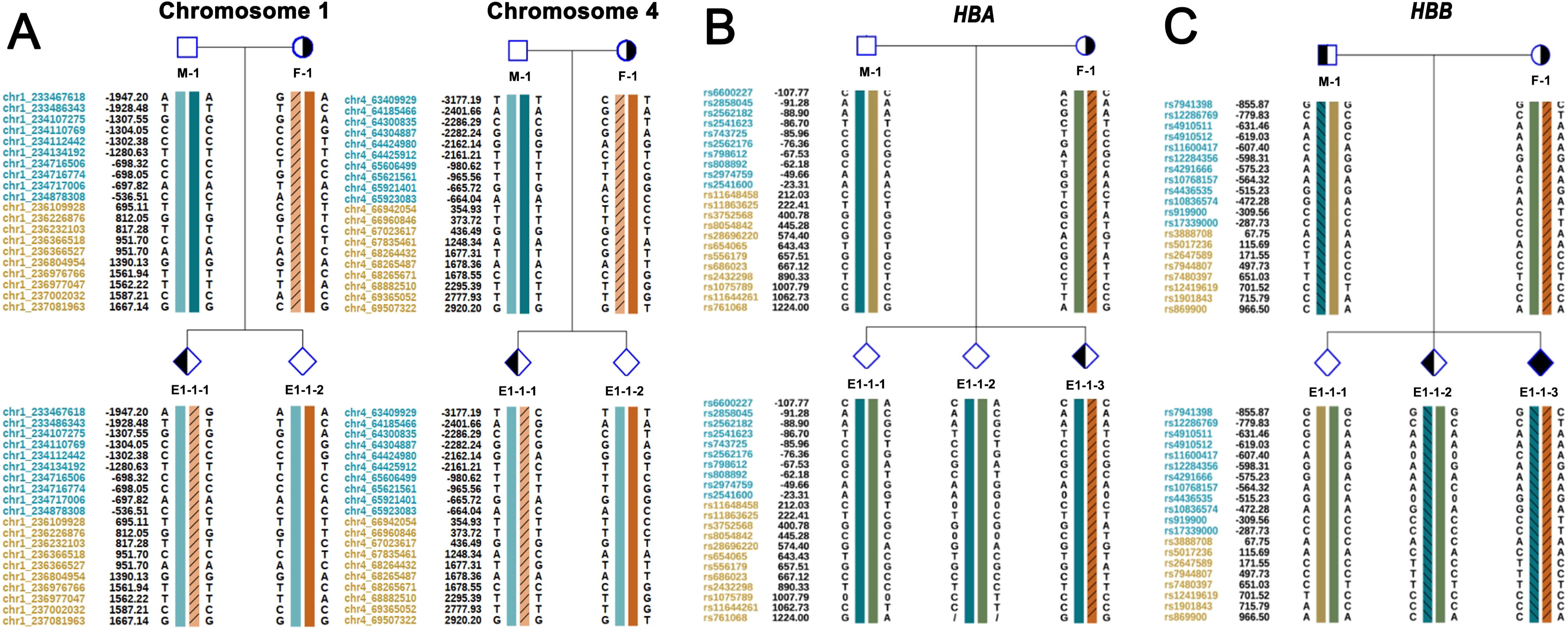
The results of haplotype linkage analysis of Family 1 based on nanopore-based adaptive sampling targeted nanopore sequencing. Panel (**A**) shows the haplotype linkage analysis based on the breakpoints of translocation in chromosome 1 and chromosome 4. Panel (**B**) displays the haplotype linkage analysis according to the *HBA* gene. Panel (**C**) presents the haplotype linkage analysis according to the *HBB* gene. ‘M-1’ represents the male individual from Family 1, while ‘F-1’ denotes the female individual from Family 1. Embryos are labeled ‘E-Family Number-Cycle Number-Embryo Number,’ e.g., ‘E1-1-1’ denotes Embryo 1 from Family 1 in the first cycle, and so forth.

There were six embryos in Family 2, including E2-2-1, E2-2-2, E2-2-3, E2-3-1, E2-4-1, and E2-4-2. All of these embryos were aneuploid, as indicated by the CNV results in Table 2.

In Family 3, seven embryos were obtained, designated E3-1-1, E3-1-2, E3-1-3, E3-1-4, E3-1-5, E3-1-6, and E3-1-7. CNV analysis revealed that E3-1-1 presented as a suspected mosaic embryo, whereas the remaining embryos displayed chromosomal abnormalities indicative of aneuploidy. These CNV findings are detailed in Table 2. Owing to the absence of euploid embryos in this cycle of ovulation promotion, we proceeded with haplotype linkage analysis on embryo E3-1-1. The results indicated that E3-1-1 harboured a maternal *HBB*:c.126_129delCTTT mutation and a paternal translocation, as depicted in Figure S5A. Additionally, Sanger sequencing was performed to validate this embryo, and the results are depicted in Figure S4B.

For Family 4, we obtained three embryos through intracytoplasmic sperm injection, designated E4-1-1, E4-1-2, and E4-2-1. Only E4-1-2 was identified as a euploid embryo, and the CNV results are provided in Table 2. Additionally, we conducted SNP detection and haplotype linkage analysis for E4-1-2. The NGS-based genotyping results are consistent with those obtained from ASTN-seq, which revealed that E4-1-2 carried both the paternal *HBB*:c.52A>T mutation and the maternal *HBB*:c.126_129delCTTT mutation, as depicted in Figure S5B. Furthermore, Sanger sequencing verification indicated that E4-1-2 harboured both *HBB*:c.52A>T and *HBB*:c.126_129delCTTT mutations (Figure S4C).

Family 5 had nine embryos, designated E5-1-1, E5-1-2, E5-1-3, E5-1-4, E5-1-5, E5-1-6, E5-1-7, E5-1-8, and E5-1-9. CNV analysis using NGS revealed that E5-1-1, E5-1-3, and E5-1-8 were euploid embryos, as shown in Table 2. Further haplotype linkage analysis based on SNPs obtained from NGS revealed that both E5-1-1 and E5-1-3 carried the maternal translocation, the maternal *HBB*:c.-43C>T mutation, and the paternal *HBB*:c.84_85insC mutation. E5-1-8 carried the maternal translocation and paternal *HBB*:c.84_85insC mutation but did not exhibit the maternal *HBB*:c.-43C>T mutation (Figure S5C). The results of haplotype linkage analysis based on ASTN-seq were consistent with those based on NGS, and the accuracy of the ASTN-seq findings was further confirmed by Sanger sequencing (Figure S4D).

Family 6 had 2 embryos, among which E6-1-2 was euploid, but the linkage analysis results indicated that both embryos carried the paternal *PKD1* gene mutation. Family 7 had a total of 10 embryos, among which E7-1-3, E7-1-5, E7-1-6, E7-1-8, and E7-1-10 were euploid. Linkage analysis indicated that E7-1-3 and E7-1-10 carried the maternal *ASNS* gene mutation but not the paternal *ASNS* gene mutation, whereas E7-1-5 and E7-1-6 carried the paternal *ASNS* gene mutation but not the maternal *ASNS* gene mutation. E7-1-8 carried both the maternal and paternal *ASNS* gene mutations. Family 8 had a total of five embryos, among which E8-1-1, E8-1-2, and E8-1-5 were euploid. Linkage analysis revealed that E8-1-1 and E8-1-2 carried both the paternal and maternal *ALPL* gene mutations, whereas E8-1-5 did not carry either the paternal or maternal *ALPL* gene mutations. The detailed results of the CNV and linkage analyses are presented in Table 2 and Figure S6, respectively. The linkage analysis results were validated by Sanger sequencing and were consistent with the results obtained from the NGS-based linkage analysis.

The results of the haplotype linkage analysis based on ASTN-seq were consistent with those of NGS, indicating that ASTN-seq is accurate and reliable. ASTN-seq enables simultaneous targeted sequencing of multiple genomic regions, making it suitable for PGT-M and PGT-SR. We can even perform linkage analysis for both monogenic disorders and chromosomal rearrangements in a single experiment.

### Embryo transfer and prenatal diagnosis

Three families (Family 1, Family 3, and Family 5) achieved successful embryo transfer, and subsequent prenatal diagnosis verified the accuracy of the embryo assessment. Following four cycles of ART, a total of 8 embryos from Family 2 underwent diagnostic evaluation, yet none were deemed fit for transplantation. Similarly, after completing two ART cycles, Family 4 submitted a total of 3 embryos for testing, but none met the criteria for suitability for transplantation. Currently, no embryos are available for transfer in Family 6, whereas Families 7 and 8 are preparing for embryo transfer.

In Family 1, the E1-1-2 embryo was implanted, resulting in the birth of a healthy infant. Prenatal diagnosis revealed a normal karyotype but detected a mutation in the *HBB* gene. The child is currently within the age range of 0–5 years and is exhibiting normal development. In Family 3, embryo E3-1-1 was transferred. Prenatal diagnosis via amniotic fluid testing revealed the presence of a maternal *HBB*:c.126_129delCTTT deletion. Furthermore, karyotyping revealed a carrier status for balanced translocations involving chromosomes 11 and 22, which were inherited from the father. The child from Family 3 is also currently within the age range of 0–5 years, exhibiting normal development in all aspects. The transfer of embryo E5-1-8 in Family 5 ended in a miscarriage. Examination of villus tissue revealed that the embryo presented normal CNVs but inherited a maternal balanced translocation and a paternal mutation of *HBB*:c.84_85insC. The posttransplantation test results of the aforementioned three embryos were consistent with the PGT results obtained from NGS and ASTN-seq (Table 2).

## Discussion

In this study, we employed NGS and ASTN-seq to perform PGT-M and PGT-SR, respectively [22]. Eight families were included in the study, *HBB* of which had at least one partner carrying both a monogenic disorder and a balanced translocation, whereas three had at least one partner carrying a monogenic disease. Genomic regions containing pathogenic mutations and translocation breakpoints were identified on the basis of chromosome karyotyping and reports from single-gene disease diagnoses. Adaptive sequencing mode was used to conduct targeted nanopore sequencing of the ROI without additional experimental operations. Through ASTN-seq, we successfully detected various mutations and performed genotyping based on flanking SNPs within a 1 Mb range around the variants. In addition, haplotype linkage analysis was conducted on euploid embryos to confirm whether they carried balanced translocations and pathogenic mutations. The accuracy of ASTN-seq-based PGT was validated with NGS-based PGT, Sanger sequencing, and prenatal diagnosis. This study represents the first application of targeted TGS in assisted reproduction, demonstrating the feasibility of ASTN-seq capable of performing both PGT-M and PGT-SR.

Researchers have developed several methods to achieve target sequencing, including multiplex PCR [23–27], probe hybridization capture [28], and CRISPR-based approaches [29–36]. However, these methods require the design and validation of primers or probes. These methods are complex to perform, lack broad applicability owing to their restriction to specific genomic loci, and result in additional costs. ASTN-seq, which is based on characteristic signal contrast has several advantages over conventional multiplex PCR, probe hybridization capture, and CRISPR. First, ASTN-seq offers a distinctive advantage in that it eliminates the need for primer or probe design and synthesis. It relies entirely on dry experimental methods, employing sequence signal comparison and sequencing voltage pulse conversion to accomplish target region enrichment. Consequently, it circumvents the complexities associated with experimental manipulation and validation. Second, ASTN-seq boasts adaptability to any genomic region, requiring only modification of the provided target sequence file. Its utilization of adaptive sampling technology comes at no extra expense, rendering it a cost-effective targeted sequencing technology. Third, ASTN-seq retains the long-read advantages of TGS.

As a newly emerging technology, ASTN-seq has been applied in mitochondrial genome sequencing [37] and direct RNA sequencing [38] because of its technological advantages. Regrettably, there are no published research papers demonstrating the application of ASTN-seq in clinical scenarios such as PGT, mutation detection, or the diagnosis of genetic diseases. Of course, other targeted TGS technologies have been used in a substantial number of articles published in the field of genetic disease detection. Methods based on multiple long-fragment PCR and TGS have been applied to genetic testing for complex inherited diseases, such as congenital adrenal hyperplasia [39], spinal muscular atrophy [40], and fragile X syndrome [41]. From a technical standpoint, ASTN-seq has an advantage in terms of read length compared with other targeted sequencing methods, but the accuracy of nanopore sequencing is not as high as that of PacBio SMRT-seq. The future application of ASTN-seq in genetic testing is worthy of exploration.

Importantly, ASTN-seq maintains performance consistency in terms of read length and sequencing accuracy with whole-genome third-generation sequencing. The primary difference between ASTN-seq (a targeted sequencing method) and TGS (inherently a whole-genome sequencing method) is that ASTN-seq offers significantly lower sequencing costs. Below is a comprehensive and integrated technical comparison of PGT based on NGS, TGS, and ASTN-seq. First, we compared the results of haplotype linkage analysis using ASTN-seq-based PGT and NGS-based PGT and found consistent results, indicating the accuracy and reliability of the ASTN-seq-based genotyping approach. Second, ASTN-seq-based PGT can construct haplotypes without requiring prior knowledge of probands such as TGS-based PGT, making it suitable for clinical scenarios where family genetic information is incomplete or involves *de novo* mutations. The read length of ASTN-seq and TGS is long enough to include pathogenic variants and SNPs in the same sequencing reads, generating natural haplotypes. Third, compared with TGS-based WGS, ASTN-seq can reduce the cost from approximately $1,100-1,200 to approximately $500-600 while maintaining the same sequencing depth of 30× for the ROI. The cost reduction caused by ASTN-seq will promote the practical application of TGS in clinical PGT. Fourth, compared with other targeted sequencing technologies, ASTN-seq possesses unique advantages while maintaining the long-read benefits of TGS. ASTN-seq is a completely dry-laboratory method that does not require the design and synthesis of primers or probes. The flexible nature of ASTN-seq makes it applicable to any genomic location and capable of detecting various types of variants. Thus, ASTN-seq can address PGT-M and PGT-SR simultaneously, avoiding multiple complex tests. Our ASTN-seq approach provides a reliable, cost-effective, and flexible solution for PGT, offering valuable references for the clinical promotion of TGS.

However, ASTN-seq has certain limitations in terms of enrichment efficiency. As shown in Figure 2A, the mean and median values of enrichment efficiency were 6.96% and 6.69%, respectively, indicating that only approximately 7 Gb of sequencing data were from the target regions among 100 Gb of total sequencing data. This relatively low enrichment efficiency is related to the technical principles of ASTN-seq. ASTN-seq sequences all nucleic acid molecules passing through the nanopore for a while before determining whether they belong to the target sequence or not. This targeting principle results in a significant waste of data, leading to reduced enrichment efficiency. We are exploring methods to improve the enrichment efficiency of ASTN-seq, such as using a combination of dry and wet experimental targeting, where initial enrichment of the target genomic region of the sample is performed through PCR amplification or probe hybridization before proceeding with ASTN-seq. We estimate that this dual-targeting method can significantly improve the enrichment efficiency of ASTN-seq from ∼6% to ∼50%.

With the continuous improvement of ASTN-seq, it holds promise for achieving the so-called “PGT-All” in the future. PGT-All refers to performing PGT-A, PGT-SR, and PGT-M simultaneously on WGA products from embryo biopsy cells with a single experimental workflow. PGT-All detects CNVs, SNVs, InDels, and SVs. It also enables the construction of haplotypes based on SNPs. The critical advantage of this technique lies in its independence from probands and the genetic information of pedigrees. A single ASTN-seq sequencing procedure is adequate for the simultaneous completion of PGT-A, PGT-SR, and PGT-M analyses. Particularly beneficial for couples harbouring multiple variants, this cost-effective and flexible approach serves to conserve substantial time and reduce testing expenditures. ASTN-seq-based PGT-All represents an ideal form of PGT technology, although much research is still needed to achieve this goal.

## Ethics statement

The procedures performed in this study and data collection were approved by the ethics committee of the Sixth Affiliated Hospital of Sun Yat-Sen University (Program No.2023ZSLYFEC-002).

## Data availability

The raw sequence data reported in this paper have been deposited in the Genome Sequence Archive (Genomics, Proteomics & Bioinformatics 2021) in National Genomics Data Center (Nucleic Acids Res 2024), China National Center for Bioinformation / Beijing Institute of Genomics, Chinese Academy of Sciences (GSA-Human: HRA009115) that are publicly accessible at https://ngdc.cncb.ac.cn/gsa-human.

## CRediT author statement

**Zhiqiang Zhang**: Data curation, Validation, Writing – original draft. **Shujing He**: Investigation, Writing – original draft. **Taoli Ding**: Methodology, Writing – original draft. **Xiaoyan Liang**: Conceptualization, Resources, Supervision. **Cong Fang**: Resources, Supervision. **Haitao Zeng**: Resources, Supervision. **Linan Xu**: Conceptualization, Formal analysis. **Xiaolan Li**: Conceptualization, Formal analysis. **Lei Jia**: Methodology, Validation. **Shihui Zhang**: Methodology, Validation. **Wenlong Su**: Methodology, Validation. **Peng Sun**: Methodology, Validation. **Ji Yang**: Data curation, Software, Visualization. **Jun Ren**: Data curation, Software, Visualization. **Sijia Lu**: Project administration, Writing – review & editing. **Zi Ren**: Project administration, Writing – review & editing.

## Acknowledgements

This work was supported by the National Key Research and Development Program of China (Grant No. 2021YFC2700503 and Grant No. 2022YFC2703200) and the National Natural Science Foundation of China (Grant No. 82271651). We would like to thank all of the investigators who participated in the present study. We also acknowledge that this paper was edited for English language, grammar, punctuation, spelling, and style by one or more highly qualified English-speaking editors at AJE.

## Supplementary material

**Figure S1 The technical flowchart of preimplantation genetic testing based on ASTN-seq**

ASTN-seq: nanopore-based adaptive sampling targeted nanopore sequencing; SNPs: single nucleotide polymorphisms; ART: assisted reproductive technology.

**Figure S2 Verification of the accuracy of translocation breakpoints detected by ASTN-seq using primer amplification**

The black text beneath the electrophoresis diagram indicates the names of the primers used, while the white text above the electrophoresis bands denotes the samples corresponding to each band. PC represents gDNA from a normal individual, and NC serves as the blank control. Each individual is labeled as ‘Gender-Family Number,’ e.g., ‘M-1’ for the male and ‘F-1’ for the female in Family 1, with the same format applied to other families. Electrophoresis band diagrams: (**A**) Family 1; (**B**) Family 2; (**C**) Family 4; (**D**) Family 5; (**E**) Family 3, where verification failed due to challenges in primer design.

**Figure S3 The IGVs of the variants**

**A.** Family 6 (M-6, *PKD1*:c.1418_1419del). **B**. Family 7 (M-7, *ASNS*:c.1193A>C; F-7, *ASNS*:c.1281delC). **C**. Family 8 (M-8, *ALPL*:c.707A>G; F-8, *ALPL*:c.407G>A).

**Figure S4 Verification of *HBB* gene mutations in euploid and mosaic embryos by Sanger sequencing**

**A.** Sanger verification diagram of the *HBB*:c.126_129delCTTT mutation in two euploid embryos, E1-1-1 and E1-1-2, in Family 1. **B**. Sanger verification diagram of *HBB* gene *HBB*:c.316-197C>T and *HBB*:c.126_129delCTTT mutations in the E3-1-1 mosaic embryo in Family 3. **C**. Sanger verification diagram of *HBB*:c.52A>T and *HBB*:c.126_129delCTTT mutations in the E4-1-2 euploid embryo in Family 4. **D**. Sanger verification diagram of *HBB*:c.-43C>T and *HBB*:c.84_85insC mutations in three euploid embryos, E5-1-1, E5-1-3, and E5-1-8, in Family 5. Embryos are named ‘E-Family Number-Cycle Number-Embryo Number,’ e.g., ‘E1-1-1’ for Embryo 1 from Family 1 in the first cycle, and similarly for others.

**Figure S5 Family haplotype linkage analysis results based on nanopore-based adaptive sampling targeted nanopore sequencing**

**A.** Haplotype linkage analysis was performed on the basis of the translocation breakpoints of chromosomes 11 and 22 in Family 3, and haplotype linkage analysis was performed on the basis of the *HBB* gene. **B**. Haplotype linkage analysis based on the translocation breakpoints of chromosomes 2 and 6 in Family 4 and haplotype linkage analysis based on the *HBB* gene. **C**. Haplotype linkage analysis based on the translocation breakpoints of chromosome 4 and chromosome 6 in Family 5 and haplotype linkage analysis based on the *HBB* gene. Individuals are labeled ‘Gender-Family Number’ (e.g., ‘M-1’ for the male and ‘F-1’ for the female in Family 1). Embryos follow the format ‘E-Family Number-Cycle Number-Embryo Number’ (e.g., ‘E1-1-1’ for Embryo 1 from Family 1 in the first cycle).

**Figure S6 Haplotype linkage analysis results based on ASTN-seq**

**A.** Family 6. **B**. Family 7. **C**. Family 8. Individuals are designated as ‘Gender-Family Number’ (e.g., ‘M-1’ for the male and ‘F-1’ for the female in Family 1). Embryos are labeled ‘E-Family Number-Cycle Number-Embryo Number’ (e.g., ‘E1-1-1’ for Embryo 1 from Family 1 in the first cycle).

## References

[1] Rubio C, Simón C. Embryo genetics. Genes 2021; 12: 118.

[2] Chow JF, Yeung WS, Lee VC, Lau EY, Ng EH. Evaluation of preimplantation genetic testing for chromosomal structural rearrangement by a commonly used next generation sequencing workflow. Eur J Obstet Gynecol Reprod Biol 2018;224:66–73.

[3] Priya PK, Mishra VV, Roy P, Patel H. A study on balanced chromosomal translocations in couples with recurrent pregnancy loss. J Hum Reprod Sci 2018;11:337.

[4] Dong B, Chen B, Liang Q, He S, Lyu W, Liu B, et al. Study on the characteristics of major birth defects in 1.69 million cases of fetus in Guangxi Zhuang Autonomous Region. Zhonghua liu Xing Bing xue za zhi 2019;40:1554–9.

[5] Lai K, Huang G, Su L, He Y. The prevalence of thalassemia in mainland China: evidence from epidemiological surveys. Sci Rep 2017;7:920.

[6] Doroftei B, Ilie O-D, Anton N, Armeanu T, Ilea C. A mini-review regarding the clinical outcomes of in vitro fertilization (IVF) following pre-implantation genetic testing (PGT)-next generation sequencing (NGS) approach. Diagnostics 2022;12:1911.

[7] Chow JF, Cheng HH, Lau EY, Yeung WS, Ng EH. Distinguishing between carrier and noncarrier embryos with the use of long-read sequencing in preimplantation genetic testing for reciprocal translocations. Genomics 2020;112:494–500.

[8] Miao H, Zhou J, Yang Q, Liang F, Wang D, Ma N, et al. Long-read sequencing identified a causal structural variant in an exome-negative case and enabled preimplantation genetic diagnosis. Hereditas 2018;155:1–9.

[9] Zhang S, Liang F, Lei C, Wu J, Fu J, Yang Q, et al. Long-read sequencing and haplotype linkage analysis enabled preimplantation genetic testing for patients carrying pathogenic inversions. J Med Genet 2019;56:741–9.

[10] Wei S, Weiss ZR, Gaur P, Forman E, Williams Z. Rapid preimplantation genetic screening using a handheld, nanopore-based DNA sequencer. Fertil Steril 2018;110:910–6.e2.

[11] Liu S, Wang H, Leigh D, Cram DS, Wang L, Yao Y. Third-generation sequencing: any future opportunities for PGT? J Assist Reprod Genet 2021;38:357–64.

[12] Chow JF, Cheng HH, Lau EY, Yeung WS, Ng EH. High-resolution mapping of reciprocal translocation breakpoints using long-read sequencing. MethodsX 2019;6:2499–503.

[13] MM YC, Yu Q, Ma M, Wang H, Tian S, Zhang W, et al. Variant haplophasing by long-read sequencing: a new approach to preimplantation genetic testing workups. Fertil Steril 2021;116:774–83.

[14] Xia Q, Li S, Ding T, Liu Z, Liu J, Li Y, et al. Nanopore sequencing for detecting reciprocal translocation carrier status in preimplantation genetic testing. BMC genomics 2023;24:1.

[15] Peng C, Chen H, Ren J, Zhou F, Li Y, Keqie Y, et al. A long-read sequencing and SNP haplotype-based novel preimplantation genetic testing method for female ADPKD patient with de novo PKD1 mutation. BMC Genomics 2023;24:521.

[16] Lu H, Giordano F, Ning Z. Oxford Nanopore MinION sequencing and genome assembly. Genomics, proteomics & bioinformatics 2016;14:265–79.

[17] Gardner DK, Lane M, Stevens J, Schlenker T, Schoolcraft WB. Blastocyst score affects implantation and pregnancy outcome: towards a single blastocyst transfer. Fertil Steril 2000;73:1155–8.

[18] Liu Y, Sun X-C, Lv G-J, Liu J-H, Sun C, Mu K. Amniotic fluid karyotype analysis and prenatal diagnosis strategy of 3117 pregnant women with amniocentesis indication. J Comp Eff Res 2023;12:e220168.

[19] Wu H, Wang H, Lan L, Zeng M, Guo W, Zheng Z, et al. Invasive molecular prenatal diagnosis of alpha and beta thalassemia among Hakka pregnant women. Medicine 2018;97.

[20] Li D, Liao C, Li J, Huang Y, Xie X, Wei J, et al. Prenatal diagnosis of β-thalassemia by reverse dot-blot hybridization in southern China. Hemoglobin 2006;30:365–70.

[21] Buchovecky CM, Nahum O, Levy B. Assessment of maternal cell contamination in prenatal samples by quantitative fluorescent PCR (QF-PCR). Methods Mol Biol 2019:117–27.

[22] Xu J, Zhang Z, Niu W, Yang Q, Yao G, Shi S, et al. Mapping allele with resolved carrier status of Robertsonian and reciprocal translocation in human preimplantation embryos. Proc Natl Acad Sci USA 2017;114:E8695–E8702.

[23] Deng X, Achari A, Federman S, Yu G, Somasekar S, Bártolo I, et al. Metagenomic sequencing with spiked primer enrichment for viral diagnostics and genomic surveillance. Nat Microbiol 2020;5:443–54.

[24] Quick J, Grubaugh ND, Pullan ST, Claro IM, Smith AD, Gangavarapu K, et al. Multiplex PCR method for MinION and Illumina sequencing of Zika and other virus genomes directly from clinical samples. Nat Protoc 2017;12:1261–76.

[25] Liu H, Li J, Lin Y, Bo X, Song H, Li K, et al. Assessment of two-pool multiplex long-amplicon nanopore sequencing of SARS-CoV-2. J Med Virol 2022;94:327–34.

[26] Mackie J, Kinoti WM, Chahal SI, Lovelock DA, Campbell PR, Tran-Nguyen LT, et al. Targeted Whole Genome Sequencing (TWG-Seq) of Cucumber Green Mottle Mosaic Virus Using Tiled Amplicon Multiplex PCR and Nanopore Sequencing. Plants 2022;11:2716.

[27] Zhang C, Xiu L, Li Y, Sun L, Li Y, Zeng Y, et al. Multiplex PCR and nanopore sequencing of genes associated with antimicrobial resistance in Neisseria gonorrhoeae directly from clinical samples. Clin Chem 2021;67:610–20.

[28] Yamaguchi K, Kasajima R, Takane K, Hatakeyama S, Shimizu E, Yamaguchi R, et al. Application of targeted nanopore sequencing for the screening and determination of structural variants in patients with Lynch syndrome. J Hum Genet 2021;66:1053–60.

[29] Gilpatrick T, Lee I, Graham JE, Raimondeau E, Bowen R, Heron A, et al. Targeted nanopore sequencing with Cas9-guided adapter ligation. Nat Biotechnol 2020;38:433–8.

[30] Simpson BP, Yrigollen CM, Izda A, Davidson BL. Targeted long-read sequencing captures CRISPR editing and AAV integration outcomes in brain. Mol Ther 2023;31:760–73.

[31] Geng K, Merino LG, Wedemann L, Martens A, Sobota M, Sanchez YP, et al. Target-enriched nanopore sequencing and de novo assembly reveals co-occurrences of complex on-target genomic rearrangements induced by CRISPR-Cas9 in human cells. Genome Res 2022;32:1876–91.

[32] Lopatriello G, Maestri S, Alfano M, Papa R, Di Vittori V, De Antoni L, et al. CRISPR/Cas9-Mediated Enrichment Coupled to Nanopore Sequencing Provides a Valuable Tool for the Precise Reconstruction of Large Genomic Target Regions. Int J Mol Sci 2023;24:1076.

[33] Erdmann H, Schöberl F, Giurgiu M, Leal Silva RM, Scholz V, Scharf F, et al. Parallel in-depth analysis of repeat expansions in ataxia patients by long-read sequencing. Brain 2023;146:1831–43.

[34] Gabrieli T, Sharim H, Fridman D, Arbib N, Michaeli Y, Ebenstein Y. Selective nanopore sequencing of human BRCA1 by Cas9-assisted targeting of chromosome segments (CATCH). Nucleic Acids Res 2018;46:e87.

[35] López-Girona E, Davy MW, Albert NW, Hilario E, Smart ME, Kirk C, et al. CRISPR-Cas9 enrichment and long read sequencing for fine mapping in plants. Plant Methods 2020;16:121.

[36] Skowronek D, Pilz RA, Bonde L, Schamuhn OJ, Feldmann JL, Hoffjan S, et al. Cas9-Mediated Nanopore Sequencing Enables Precise Characterization of Structural Variants in CCM Genes. Int J Mol Sci 2022;23:15639.

[37] Kipp EJ, Lindsey LL, Milstein MS, Blanco CM, Baker JP, Faulk C, et al. Nanopore adaptive sampling for targeted mitochondrial genome sequencing and bloodmeal identification in hematophagous insects. Parasit Vectors 2023;16:68.

[38] Naarmann-de Vries IS, Gjerga E, Gandor CL, Dieterich C. Adaptive sampling for nanopore direct RNA-sequencing. RNA 2023;29:1939–49.

[39] Liu Y, Chen M, Liu J, Mao A, Teng Y, Yan H, et al. Comprehensive analysis of congenital adrenal hyperplasia using long-read sequencing. Clin Chem 2022;68:927–39.

[40] Li S, Han X, Xu Y, Chang C, Gao L, Li J, et al. Comprehensive analysis of spinal muscular atrophy: SMN1 copy number, intragenic mutation, and 2+0 carrier analysis by third-generation sequencing. J Mol Diagn 2022;24:1009–20.

[41] Liang Q, Liu Y, Liu Y, Duan R, Meng W, Zhan J, et al. Comprehensive analysis of fragile X syndrome: full characterization of the FMR1 locus by long-read sequencing. Clin Chem 2022;68:1529–40.

