## Supplementary figures and images for "Cost-effective and Flexible Preimplantation Genetic Testing (PGT) Using Adaptive Sampling-based Targeted Nanopore Sequencing (ASTN-seq)"

### Figure S1

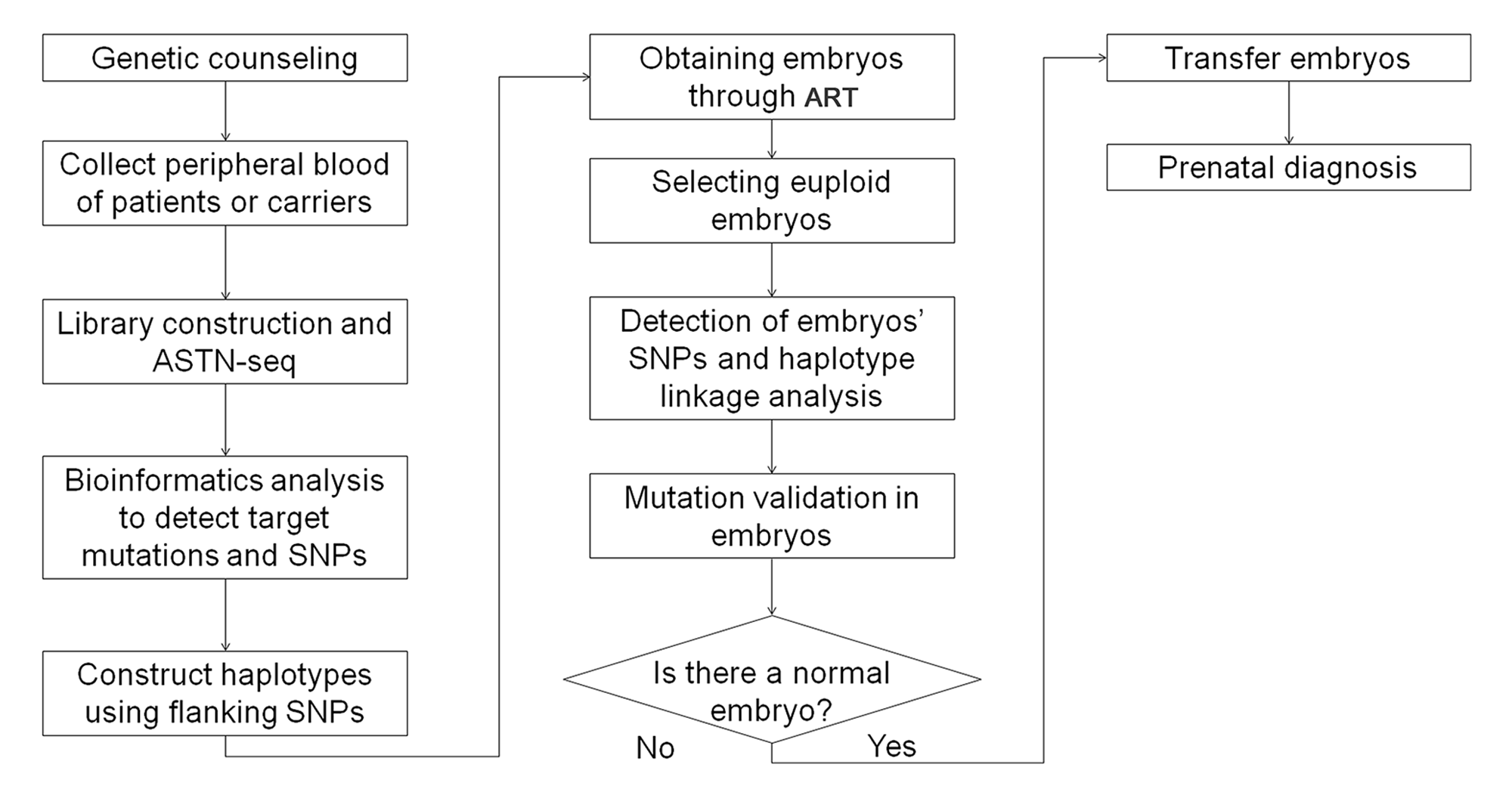

### Figure S2

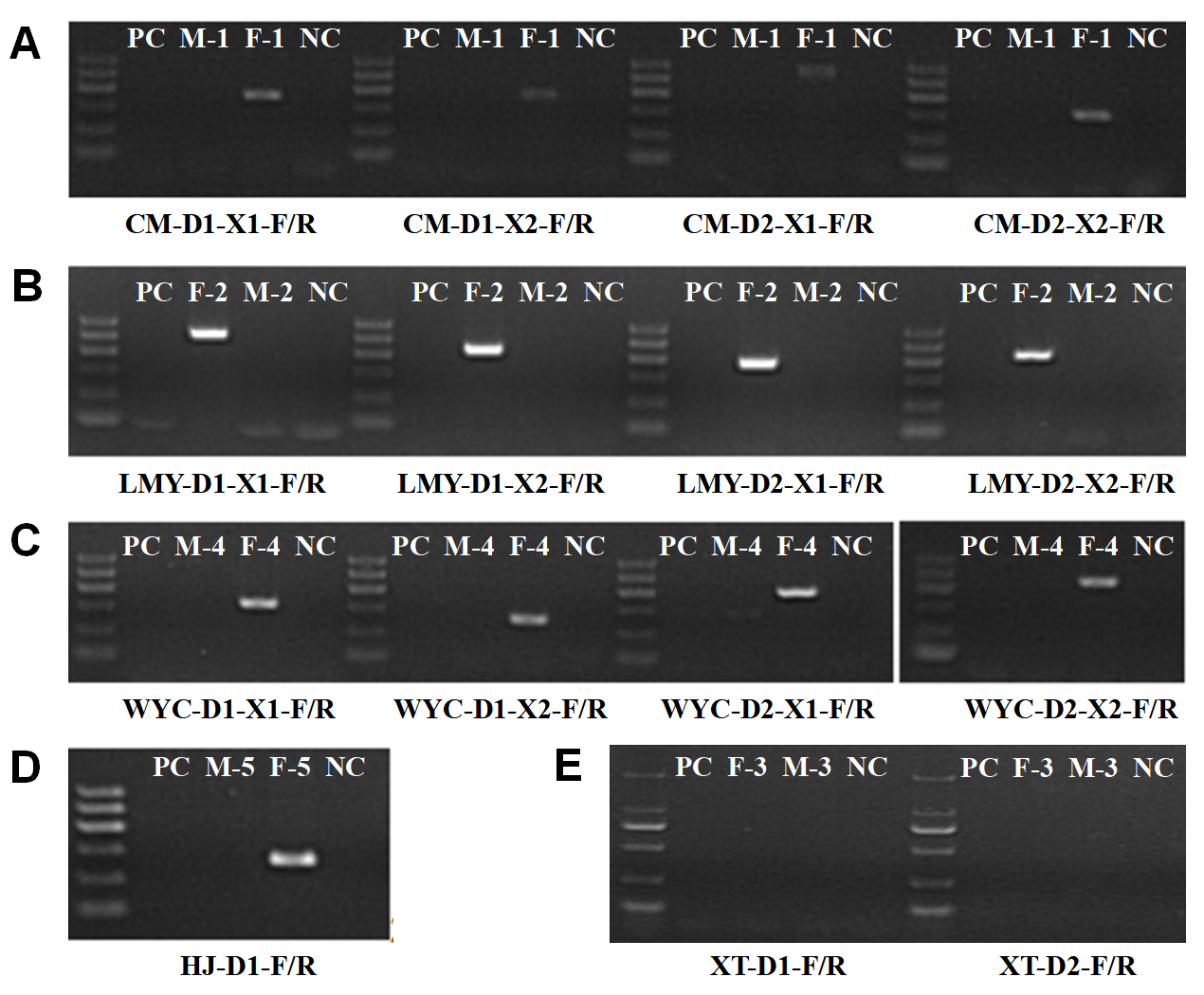

### Figure S3

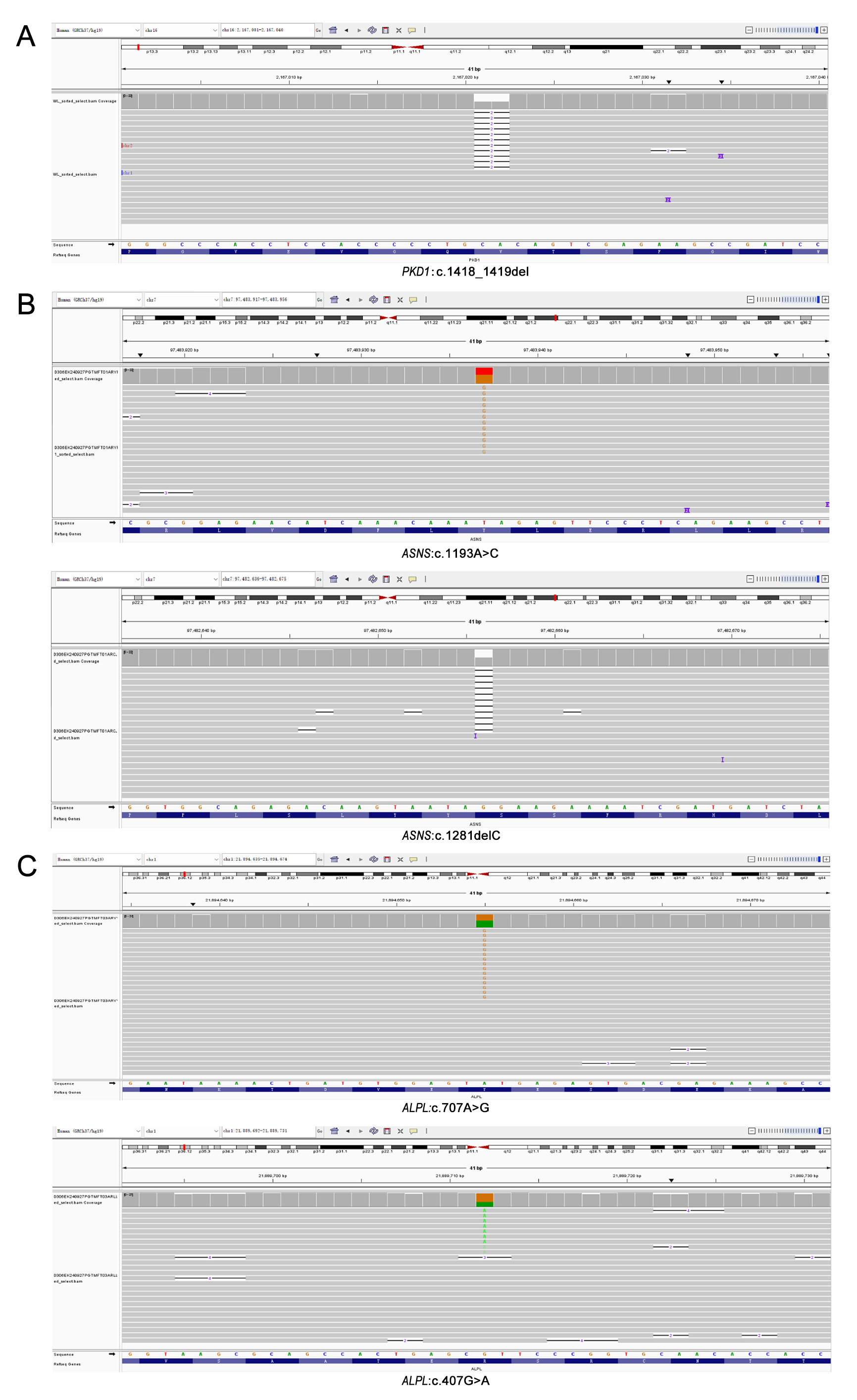

### Figure S4

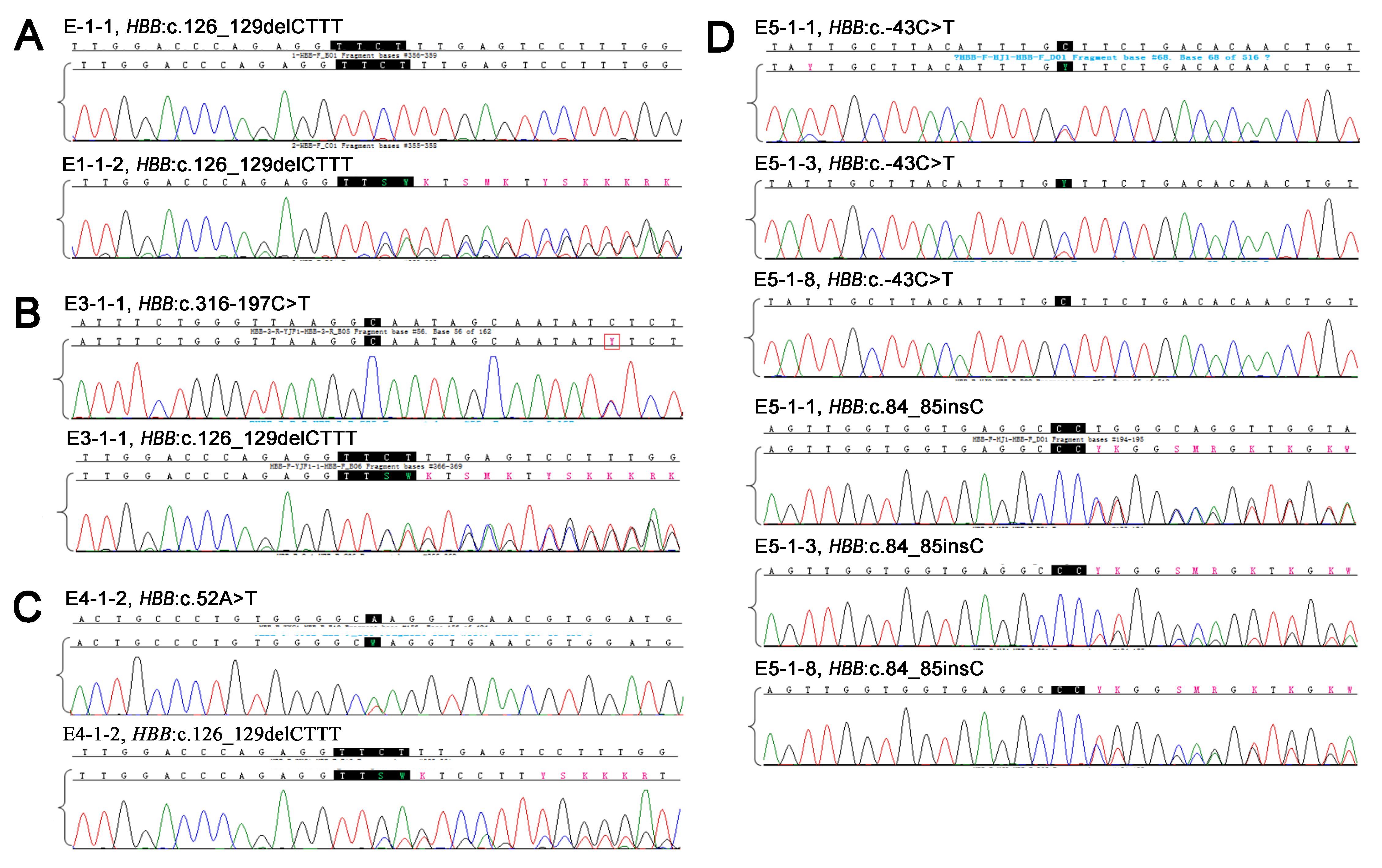

### Figure S5

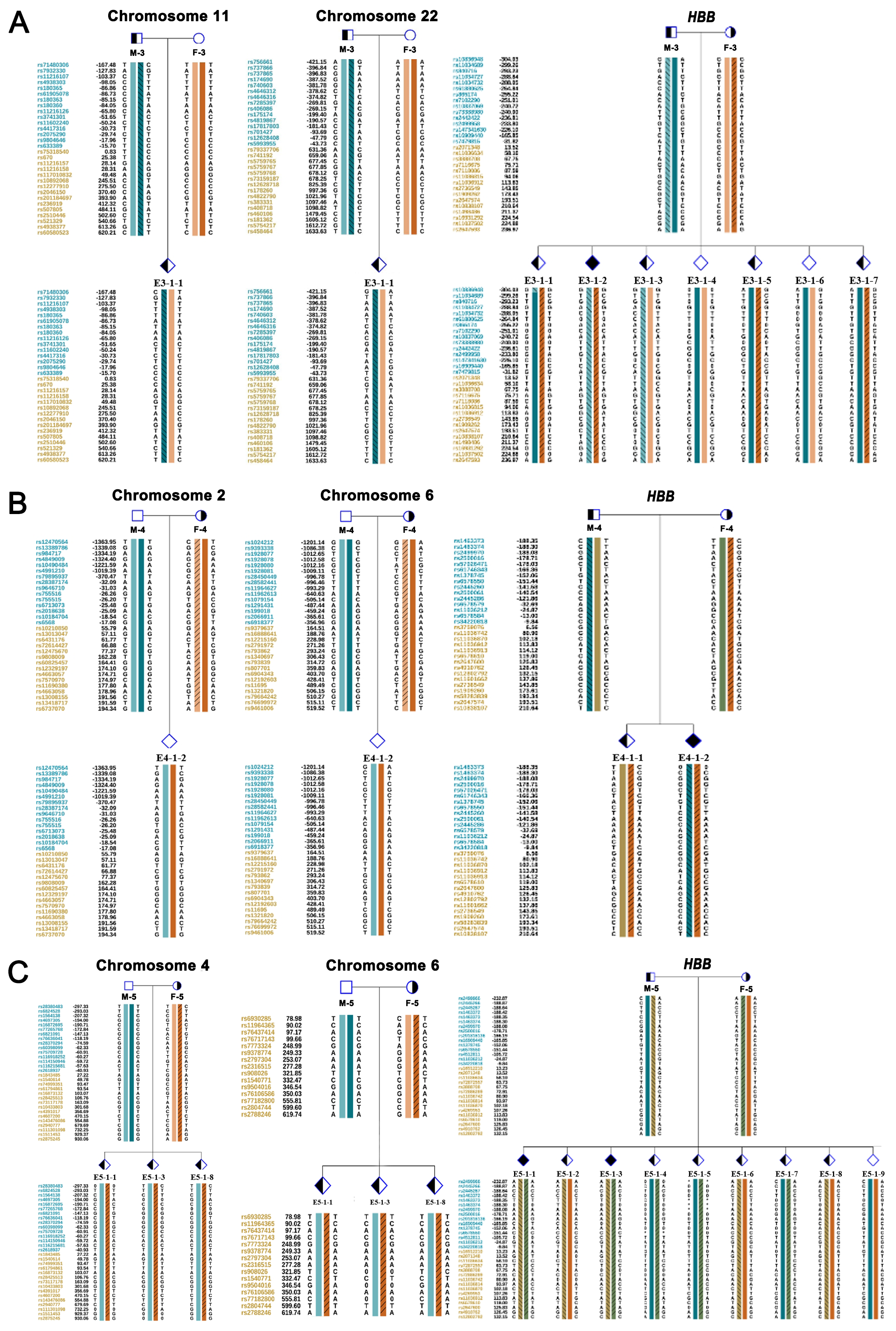

### Figure S6

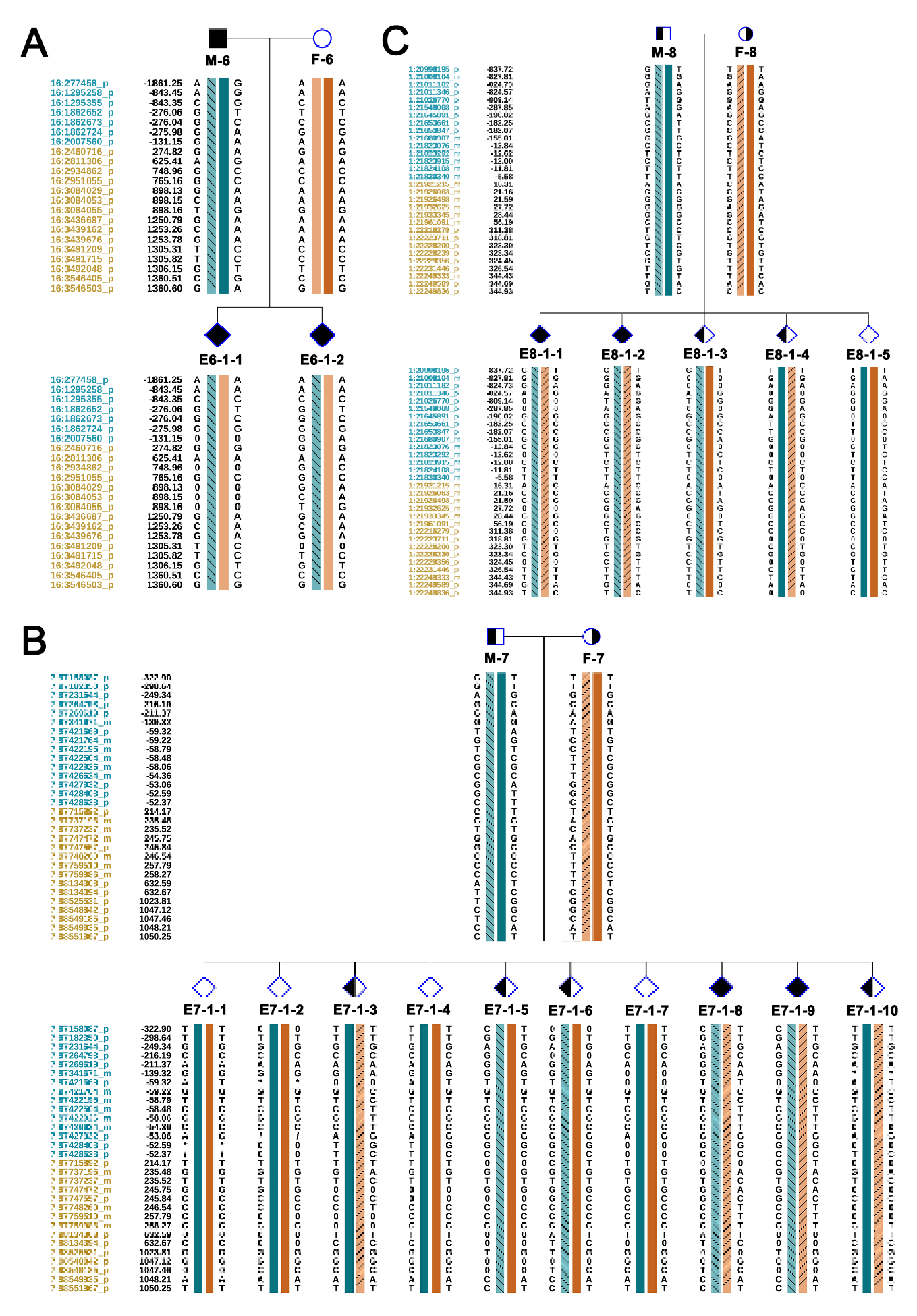
